# Adherence to Red Reflex and Vision Screening Recommendations: A Deep Dive into Primary Care Implementation Gaps

**DOI:** 10.64898/2026.06.08.26355190

**Authors:** Afua O. Asare, Giovani Robles, E. Eugenie Hartmann, Carole Stipelman, Dallen Calder, Olaoluwa Omotowa, Jessie Montgomery, Bryce T. Baugh, Brian Stagg, Guilherme Del Fiol, Melissa H. Watt, Michelle R. Hribar, J.D. Smith

## Abstract

**Introduction:** Early childhood vision screening is critical for detecting amblyopia and other vision-threatening conditions. Despite screening recommendations during well-child visits, rates remain low. Red reflex assessment is recommended to identify serious ocular pathology, yet its use in primary care is not well described. We examined rates and drivers of vision screening in pediatric primary care.

**Methods:** We conducted a retrospective review of electronic health records for children 3–5 years attending well-child visits in 2022 in one of three representative primary care clinics within a university health system. Outcomes were documented red reflex and functional vision tests. We evaluated associations with patient demographics and clinic site using multivariable logistic regression

**Results:** Among 1,003 visits, 21.1% (n=212) had a documented red reflex assessment, and 60.8% (n=610) a functional vision test. Younger children (ages 3 and 4 vs. 5 years) had higher odds of red reflex assessment [adjusted odds ratio (aOR) 9.00 and 8.64], and lower odds of a functional vision (aOR 0.47 and 0.59) test. Females had higher odds of red reflex assessment (aOR 1.53). Other/Multiracial children had lower odds of red reflex assessment than Non-Hispanic White children (aOR 0.48). Screening rates varied significantly by clinic site

**Conclusions:** Visual function and red reflex assessment are inconsistently performed in pediatric primary care, with particularly low rates of red reflex documentation. Screening rates varied between clinics and were affected by age. These findings highlight missed opportunities for early detection of vision-threatening conditions and identify targets for improving adherence to pediatric vision screening recommendations.

## Introduction

Childhood vision impairment is associated with reduced quality of life and reading difficulties, leading to poor educational outcomes.^1,2^ One in five children in the US has refractive errors or other amblyogenic risk factors that require early detection to prevent permanent visual impairment.^3–5^ For this reason, the American Academy of Pediatric Ophthalmology and Strabismus (AAPOS), the American Academy of Pediatrics (AAP), and other national associations recommend visual function tests and red reflex assessments during well-child visits.^6^ Visual function tests in pediatric primary care for children 3 to 5 years is more cost-effective than school-based screening,^7^ and more effective when delivered in a patient-centered medical home.^8^ Vision screening comprise an assessment of visual acuity using optotype charts, or refractive errors using instrument-based screening, and red reflex assessments. Following an abnormal finding in either assessment, referral to eye care professionals is recommended for diagnosis and treatment. ^6,9^ Visual function tests detect vision disorders that may cause amblyopia (amblyogenic risk factors) and refractive errors. In contrast, red reflex assessments are critical for the detection of structural abnormalities such as cataracts, glaucoma, and retinoblastoma.^9^ Although congenital cataracts and retinoblastoma are relatively rare, their potential for permanent vision loss and, in the case of retinoblastoma, mortality, underscore the necessity of early identification.^10,11^ ^12,13^

Vision screening for children is covered by most US health plans in line with the Affordable Care Act (ACA), which requires coverage for preventive care services recommended by the United States Preventive Services Taskforce with a grade of A or B level of evidence.

Reimbursement of providers for offering pediatric visual function tests is limited to commercial health plans. Medicaid in most states, including Utah, does not reimburse for pediatric visual function or red reflex assessments separate from well-child visits. A global payment for a well-child visit is provided (regardless of whether a visual function or red reflex assessment is conducted).^14,15^ Commercial plans do not reimburse providers for red reflex assessments.

Despite recommendations and the availability of insurance coverage,^16^ primary care vision screening in pediatric primary care remains low,^17^ particularly among marginalized populations and children with developmental disabilities.^18,19^ In 2019, only 27% of US children under the age of 6 received a vision screening in pediatric primary care.^17^ In a 2013 survey of primary care physicians in Ontario, Canada, 25% of respondents reported conducting a red reflex assessment in children over the age of 3 years. Prior studies assessing determinants of (‘barriers to’) evidence-based vision screening in pediatric primary care are limited by reliance on caregiver self-report,^20,21^ dated data (2013),^22^ and a narrow focus on physician and/or patient-only factors.^19,23,24^ To identify targets for interventions that bridge the gaps in access to primary care vision screening, studies are needed to understand the determinants. The objective of this study was to quantify the rate of pediatric primary care vision screening (visual function and red reflex) assessments in a single university health system and to identify patient- and contextual-level drivers that may inform future targeted interventions.

## Patients and Methods

### Study Design, Participants and Data Collection

We conducted a manual retrospective cross-sectional chart review of Epic electronic health records (EHRs) of three pediatric primary care clinics in a university health care system in Utah. We limited this study to three representative clinics to allow for manual, dual-reviewed data extraction. This approach ensured a level of data accuracy and the capture the unstructured documentation of vision screening that a large-scale, automated EHR pull would miss. We selected three clinics (A, B, and C) to represent the full range of vision screening workflows, well-child visit appointment durations, and patient populations across the entire university health care system (Supplementary Materials). All three clinics are located in Salt Lake County, an urban area that is the 2nd most racially and ethnically diverse county in Utah, with 69% of people identifying as Non-Hispanic White. ^25–27^ The university health care system consists of 30 primary care clinics, five of which serve as academic pediatric training centers. In all clinics, vision screening is performed by auxiliary medical staff, primarily medical assistants. Red reflex assessments are performed by physicians or nurse practitioners who also interpret the results to determine next steps. Workflow variations are described elsewhere (Supplement).

For this study, we reviewed EHRs of children aged 3 to 5 years who attended a well-child visit in either of the three representative pediatric primary care clinics. Between April and October 2023, we manually collected retrospective data for consecutive well-child visits beginning in January and continuing through October 2022. Details of the data abstraction process are described in Supplementary Materials. The study received an exemption from the University of Utah Institutional Review Board because it was deemed minimal risk and a review of existing EHR data. The study was designed and reported in accordance with the Strengthening the Reporting of Observational Studies in Epidemiology (STROBE) guidelines.

### Measures

The primary outcome variable was documentation of a visual function test (yes/no), which could be an assessment of visual acuity using an optotype chart, or refractive errors using an instrument-based vision screening camera [Spot Vision Screener (Welch Allyn, Skaneateles Falls, NY, United States)]. The secondary outcome variable was receipt of a red reflex assessment (yes/no). Recommended vision screening tests for children in pediatric primary care, per AAPOS and AAP, are described in the Supplementary Materials.

We selected exposure variables (contextual and patient factors) based on Andersen’s Behavioural Model of Health Services Use and assessed as categorical variables. The contextual variable was the specific pediatric primary care clinic where vision was assessed (‘clinic site’), labeled A, B, or C for anonymity. Patient factors were enabling factors [type of health insurance coverage (Medicaid/government, self-pay, commercial/private)], and predisposing factors [race and ethnicity (Hispanic/Latino, Black/Non-Hispanic, Other/Multiracial, White/Non-Hispanic and unknown or undisclosed), sex (male, female) and age (3, 4, or 5 years)]. These variables were selected because of their suspected influence on vision tests during well-child visits.^19–23^ We also determined well-child visit and provider volume for each clinic to assess possible clinic-level factors associated with the outcomes.

### Statistical analyses

We used Stata, v 18.0 (StataCorp, College Station, TX) to perform all analyses. We used chi-square tests to assess the association between categorical and outcome variables, and Pearson correlation coefficients to assess the association between continuous variables. We built fixed effects logistic regression models to estimate unadjusted odds ratio (OR), adjusted odds ratio (aOR), with 95% confidence interval (CI) for the association between the exposure and outcome variables. We also computed adjusted predicted probabilities (aPP) from logistic regression models using marginal standardization, providing interpretable estimates of outcome likelihood across categories and complementing odds ratios, which reflect relative associations. Cramer’s V was used to assess correlations between exposures. For model building, we excluded any two correlated variables with a Cramer’s V of 0.5 or higher from the final model to reduce multicollinearity. Each model (visual function test or red reflex assessment) included sex, age (year), race and ethnicity, type of health insurance, and clinic site. Statistical significance for all analyses was defined as *p* < .05. In a sub-analysis, we examined whether the interaction between race/ethnicity and health insurance type was associated with receipt of a vision screening test (visual function and red reflex).

## Results

The cohort included 1,003 children who received a well-child visit at one of three representative pediatric primary care clinics between January and October 2022 and were about equally distributed by sex (47% female) and age (35.6% aged 3, 32.4% aged 4, and 32.0% aged 5 years) (Table 1). Although the majority of the cohort was predominantly non-Hispanic White [42.2% (n=423)] and Hispanic/Latino [36.8% (n=369)], demographic characteristics varied significantly by clinic type. While over half of children had commercial health insurance [58.6% (n=588)], a significant proportion had Medicaid/government insurance [35.4% (n=355)], and 6.0% were self-pay (n=60). Clinic C was the most demographically distinct, serving a population that was primarily Hispanic (75.2% (n=248) and Medicaid insured [75.2% (n=248)].

**Table 1.** Descriptive characteristics of study participants in each of the three pediatric primary care clinics where children received well-child visits.

| **Patient Characteristics, n(%)** | **Total**<br>**(n=1,003)** | **Clinic A**<br>**(n=334)** | **Clinic B**<br>**(n=339)** | **Clinic C**<br>**(n=330)** | **p<sup>b</sup>** |
| --- | --- | --- | --- | --- | --- |
| **Sex** |  |  |  |  | 0.437 |
| Male | 535 (53.33) | 169 (50.60) | 183 (53.98) | 183 (55.45) |  |
| Female | 468 (46.67) | 165 (49.40) | 156 (46.02) | 147 (44.55) |  |
| **Age (year)** |  |  |  |  | 0.030 |
| 3 | 357 (35.59) | 101 (30.24) | 123 (36.28) | 133 (40.30) |  |
| 4 | 325 (32.40) | 115 (34.43) | 119 (35.10) | 91 (27.58) |  |
| 5 | 321 (32.00) | 118 (35.33) | 97 (28.61) | 106 (32.12) |  |
| **Race and ethnicity** |  |  |  |  | <.001 |
| Hispanic/Latino | 369 (36.79) | 48 (14.37) | 73 (21.53) | 248 (75.15) |  |
| Non-Hispanic Black | 30 (2.99) | 11 (3.29) | 7 (2.06) | 12 (3.64) |  |
| Other/Multiracial | 130 (12.96) | 49 (14.67) | 42 (12.39) | 39 (11.82) |  |
| Non-Hispanic White | 423 (42.17) | 204 (61.08) | 193 (56.93) | 26 (7.88) |  |
| *Unknown or undisclosed* | 51 (5.08) | 22 (6.59) | 24 (7.08) | 5 (1.52) |  |
| **Type of Health Insurance** |  |  |  |  | <.001 |
| Medicaid/Government | 355 (35.36) | 56 (16.77) | 51 (15.04) | 248 (75.15) |  |
| Self-Pay | 60 (5.98) | 7 (2.10) | 5 (1.47) | 48 (14.55) |  |
| Commercial/Private | 588 (58.62) | 271 (81.14) | 283 (83.48) | 34 (10.30) |  |

Overall, 60.8% (n=610) of children sampled received a visual function test, and 21.1% (n=212) received a red reflex assessment. Visual function tests across clinics ranged from 12.1% to 87.4%, while rates of red reflex assessments ranged from 2.1% to 46.3% (Figure 1). Overall, visual function rates increased with age, with 5-year-olds having the highest (36.1%) rate, and 3-year-olds having the lowest (33.0%) (Table 2). The alternative was true for red reflex assessments. Specifically, the rates of reflex assessments decreased with age, with 3-year-olds having the highest rate (46.2%) and 5-year-olds the lowest (8.1%). Hispanic/Latino children had the highest rate of visual function tests (42.8%), while non-Hispanic white children recorded the highest rate of red reflex assessments (62.7%). Most of the children with a documented visual function test or red reflex assessment had commercial/private insurance (45.4% and 83.5%, respectively). Clinic B had the lowest rate of visual function tests (12.1%), and the highest rate of red reflex assessment (46.3%) compared to clinics A and C.

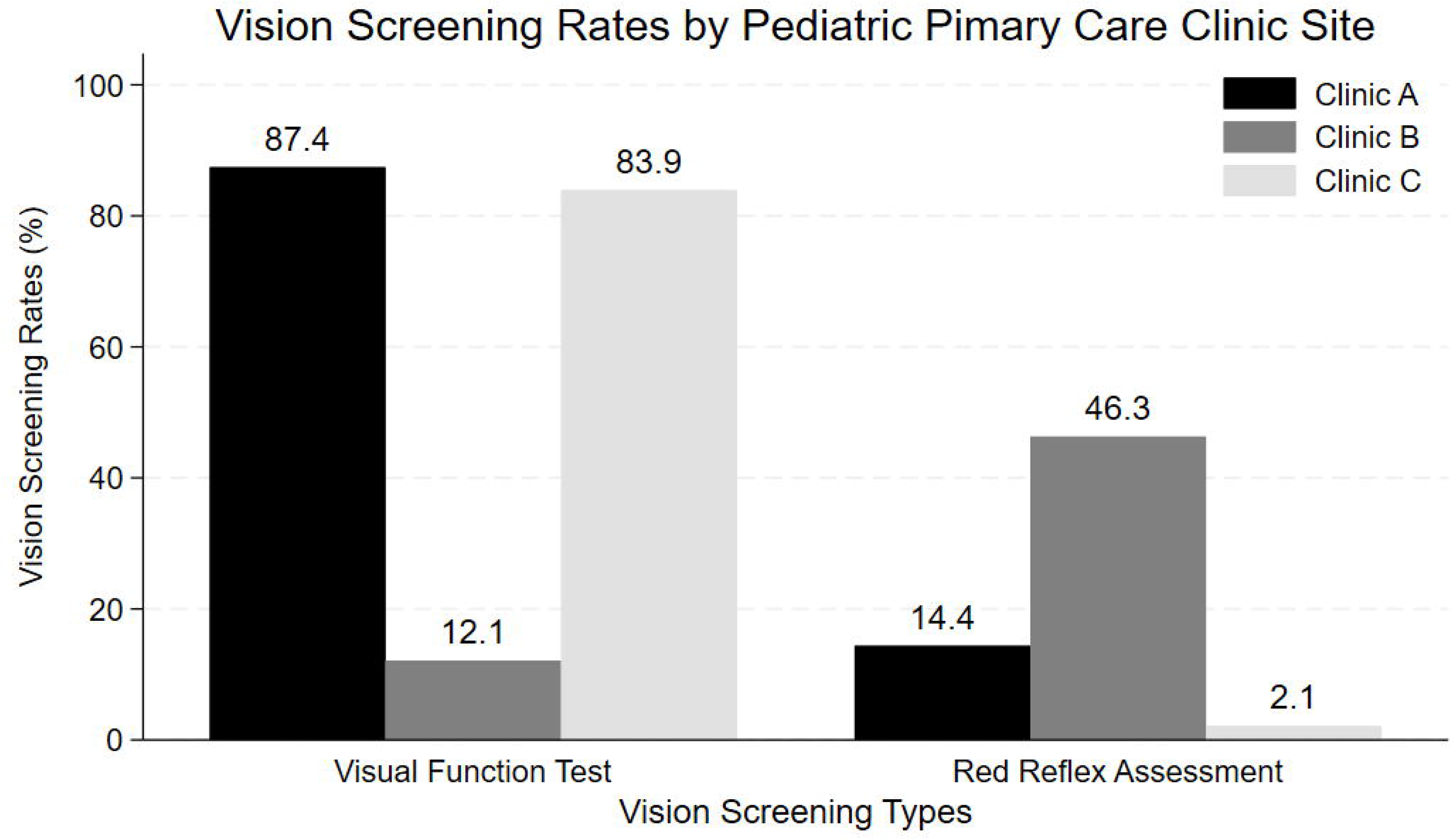

**Table 2.**
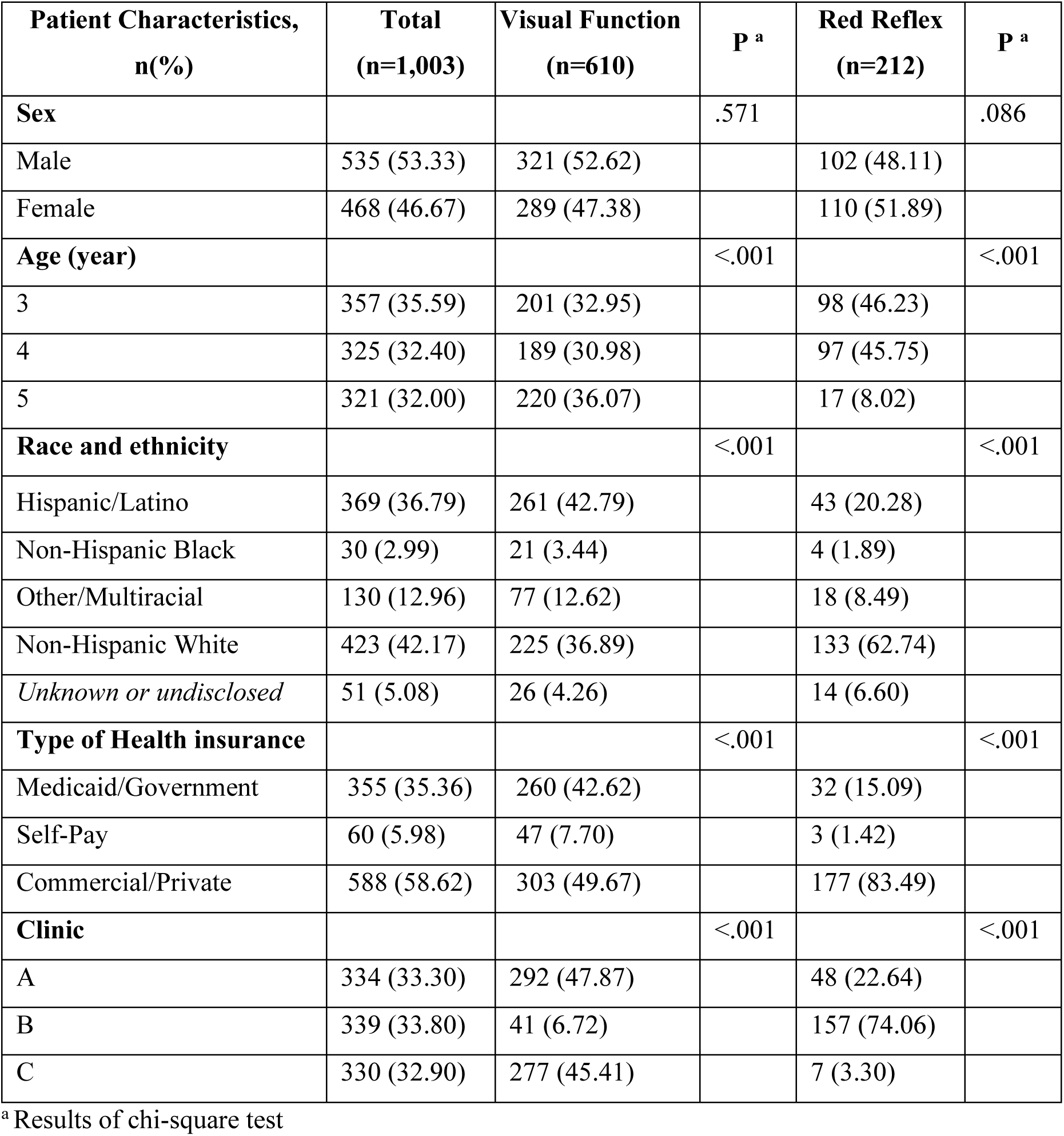
Descriptive characteristics of the number of children (%) in the cohort with a documented A) visual function test, and B) Red reflex assessments.

| **Patient Characteristics,**<br>**n(%)** | **Total**<br>**(n=1,003)** | **Visual Function**<br>**(n=610)** | **P<sup>a</sup>** | **Red Reflex**<br>**(n=212)** | **P<sup>a</sup>** |
| --- | --- | --- | --- | --- | --- |
| **Sex** |  |  | .571 |  | .086 |
| Male | 535 (53.33) | 321 (52.62) |  | 102 (48.11) |  |
| Female | 468 (46.67) | 289 (47.38) |  | 110 (51.89) |  |
| **Age (year)** |  |  | <.001 |  | <.001 |
| 3 | 357 (35.59) | 201 (32.95) |  | 98 (46.23) |  |
| 4 | 325 (32.40) | 189 (30.98) |  | 97 (45.75) |  |
| 5 | 321 (32.00) | 220 (36.07) |  | 17 (8.02) |  |
| **Race and ethnicity** |  |  | <.001 |  | <.001 |
| Hispanic/Latino | 369 (36.79) | 261 (42.79) |  | 43 (20.28) |  |
| Non-Hispanic Black | 30 (2.99) | 21 (3.44) |  | 4 (1.89) |  |
| Other/Multiracial | 130 (12.96) | 77 (12.62) |  | 18 (8.49) |  |
| Non-Hispanic White | 423 (42.17) | 225 (36.89) |  | 133 (62.74) |  |
| *Unknown or undisclosed* | 51 (5.08) | 26 (4.26) |  | 14 (6.60) |  |
| **Type of Health insurance** |  |  | <.001 |  | <.001 |
| Medicaid/Government | 355 (35.36) | 260 (42.62) |  | 32 (15.09) |  |
| Self-Pay | 60 (5.98) | 47 (7.70) |  | 3 (1.42) |  |
| Commercial/Private | 588 (58.62) | 303 (49.67) |  | 177 (83.49) |  |
| **Clinic** |  |  | <.001 |  | <.001 |
| A | 334 (33.30) | 292 (47.87) |  | 48 (22.64) |  |
| B | 339 (33.80) | 41 (6.72) |  | 157 (74.06) |  |
| C | 330 (32.90) | 277 (45.41) |  | 7 (3.30) |  |

<sup>a</sup> Results of chi-square test

Race and ethnicity were not reported for 51 children, and their data were subsequently excluded from the multivariate logistic regression analyses. After adjusting for other variables in the model, children who were 3 or 4 years old had lower odds of receiving a visual function test compared with those who were aged 5 years (adjusted odds ratio [aOR] 0.47; 95% CI 0.30, 0.76 for 3-year olds, and aOR 0.59; 95% CI 0.37, 0.96 years old for 4 year olds vs 5 year olds).

Children who were screened in Clinic B were less likely to get a visual function test compared to children in Clinic A (aOR 0.02; 95% CI 0.01, 0.03) (Table 3). Adjusted predicted probabilities and 95% CI for each exposure are described in Figure 2.

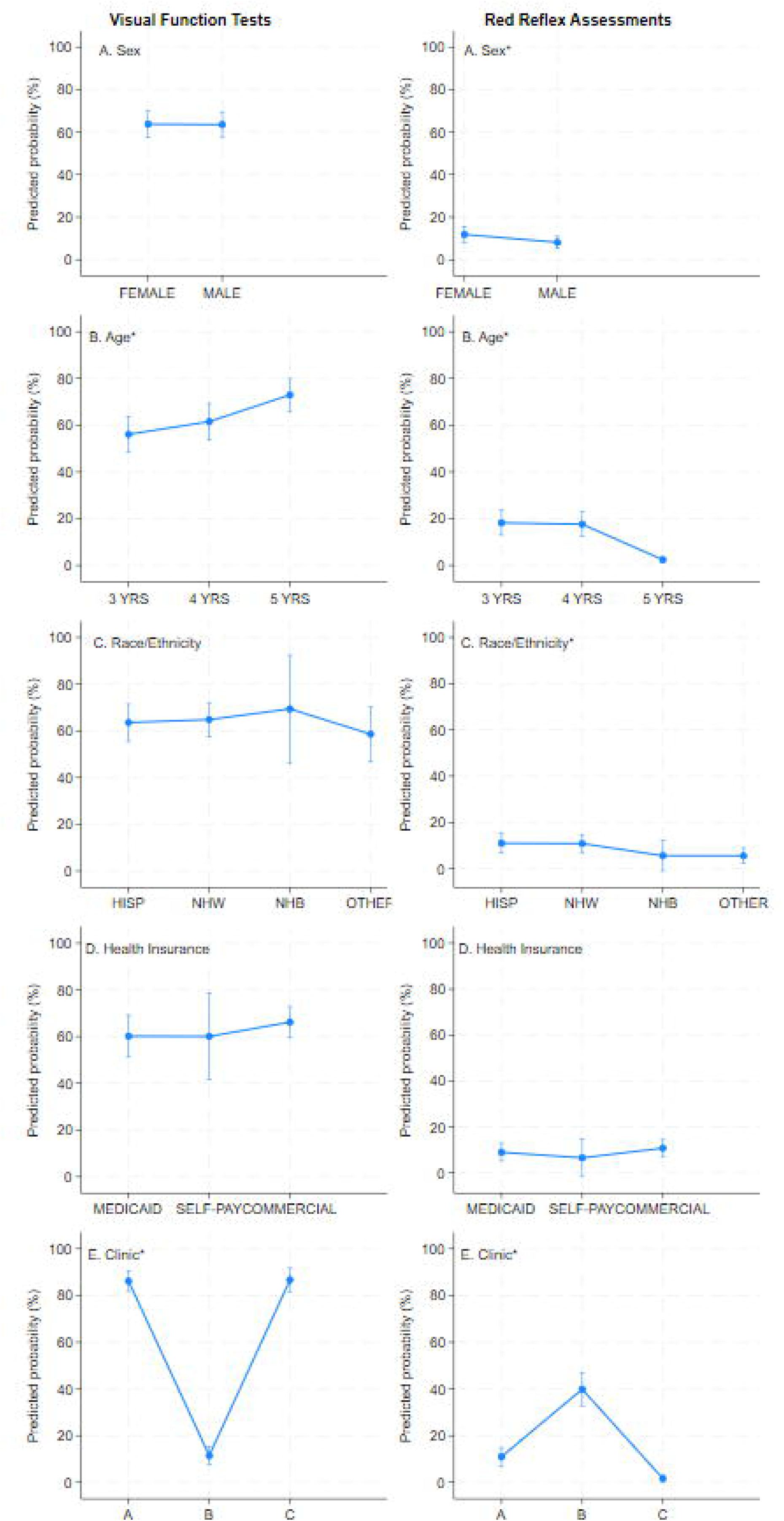

**Table 3.**
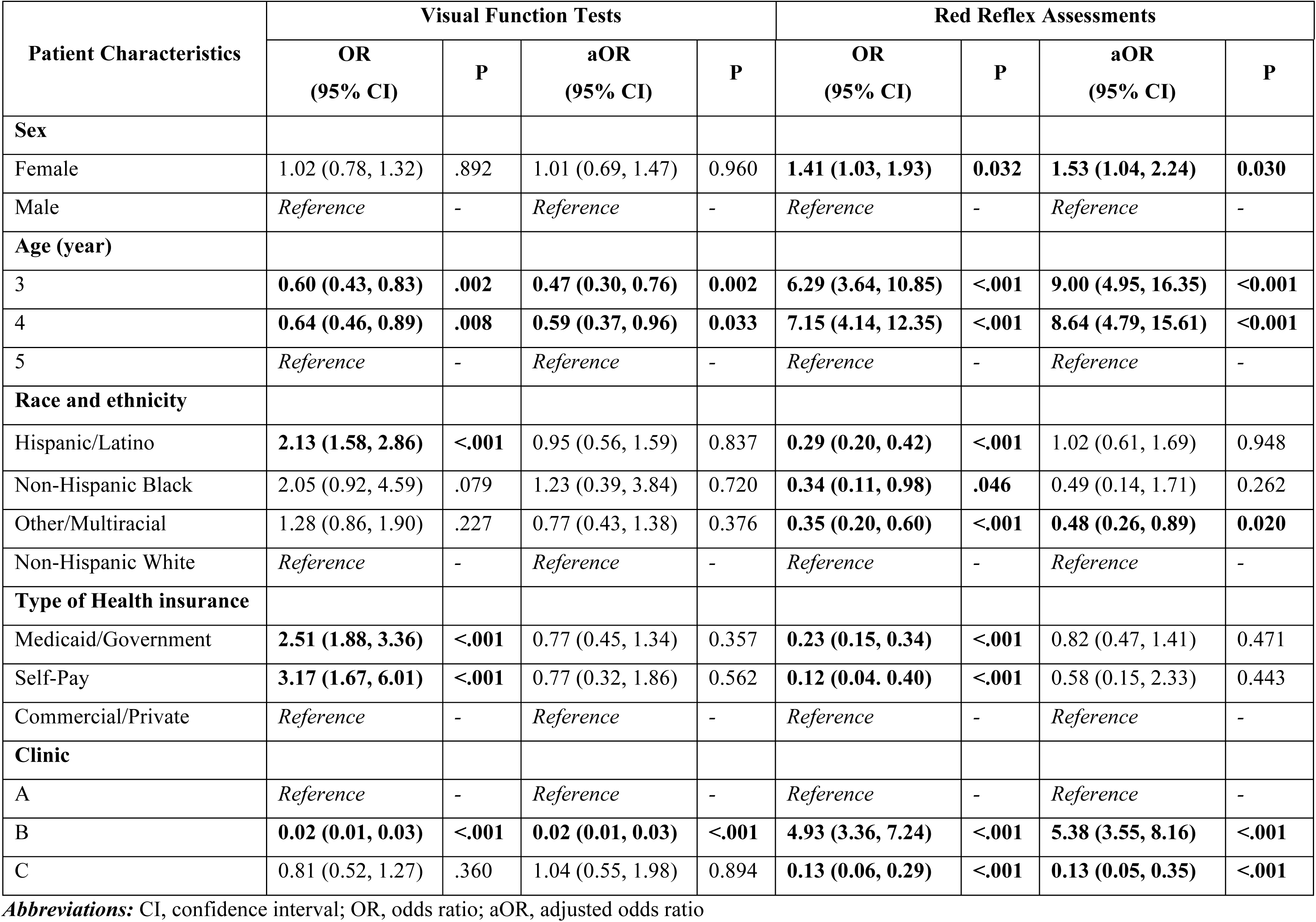
Association between contextual and individual factors, and receiving a A) visual function test (n=952), and B) red reflex assessments (n=212)

In 2022, Clinics A, B, and C recorded 846, 843, and 406 annual well-child visits, respectively. Annual well-child visit volume was negatively correlated with the number of visual function tests performed (r = -0.42, p = 0.73) and positively correlated with the number of red reflex assessments performed (r = +0.68, p = 0.52). There were 29 providers in clinic A, and 11 in clinic B who led or supported (Supplementary Materials). The number of clinic providers who conducted well-child visits was positively correlated with the number of visual function tests performed (r = 1.00, p = 0.034) and negatively correlated with the number of red reflex assessments performed (r = –0.96, p = 0.17).

Regarding receipt of red reflex assessments, after adjusting for other variables in the model, females compared to males had higher odds of having a red reflex assessment (aOR 1.51; 95% CI 1.03, 2.22) (Table 3). Children who were 3 or 4 years old had higher odds of receiving a red reflex assessment compared to children aged 5 years old [(aOR 9.02; 95% CI 4.96, 16.40) for 3-year olds, and (aOR 8.65; 95% CI 4.79, 15.61) for 4-year-olds compared to 5-year olds].

Children who identified as Other/Multiracial had a lower likelihood of undergoing a red reflex assessment than non-Hispanic white children (aOR 0.48; 95% CI 0.26, 0.89). Children who were screened in Clinic B had higher odds of getting a red reflex assessment compared to children seen in Clinic A; while those in clinic C had lower odds of getting a red reflex assessment compared to children seen in Clinic A [Clinic B vs A (aOR 5.39; 95% CI 3.55, 8.16), and Clinic C vs A (aOR 0.13; 95% CI 0.05, 0.36)]. Predicted probabilities of red reflex assessments and their 95% CI for each exposure are shown in Figure 2. In the sub-analyses assessing the association between the interaction of race and ethnicity with type of health insurance coverage and the receipt of vision tests, the associations were not statistically significant.

## Discussion

This study provided estimates of the rates of vision screening, as well as contextual and patient-level drivers, across three pediatric primary care clinics within a university health system in Utah. Importantly, the study results highlight substantial variability in pediatric vision screening rates driven by both contextual and patient-related factors, and inter-system variability. Our results demonstrate that even within a single, unified health system, the rates of vision screening function tests varied dramatically between sites (visual function 2.1% to 87.4%, and red reflex 2% to 46%). While we found that age was significantly associated with visual function and red reflex assessments, the profound variability between clinics suggests that site-specific culture and workflow are more powerful determinants of whether each screening test is even attempted. This indicates that while a child’s age may influence their ability to cooperate with a test, the specific clinic they visit ultimately determines their access to that care. Twenty-one percent of children with a well-child visit received a red reflex assessment, and 61% received a visual function test. The rate of visual function tests is consistent with that reported in a study by Hoover et al., who published a retrospective cross-sectional analysis of data from well-child visits at 40 pediatric primary care practices within Nemours Children’s Health network in Delaware, Pennsylvania (Delaware Valley), and Florida. The analysis of about 64,000 well-child visits revealed that visual function tests were completed in 59% of visits.^19^ To the best of our knowledge, no peer-reviewed studies have reported the rate of red reflex assessments amongst children in well-child visits.

Regarding contextual factors associated with receiving vision screening, clinics with high rates of visual function tests did not demonstrate high red reflex assessment, suggesting differential workflow prioritization. While prior studies have reported variability in visual function tests at the provider-level,^21,22,24,28^ no studies were identified that explored variation in visual function tests at the clinic level. Marsh-Tootle et al. reported variability in rates of visual function tests among pediatric primary care providers with high compared to low well-child visit volumes. Those with low well-child visit volume were more likely to provide visual function tests after adjusting for nesting of preschool patients within providers.^22^ In our study, clinical-level variation is identified as a strong driver of the administration of visual function tests, which is likely due to clinical culture, workflow differences, equipment availability, and lack of reimbursement. These clinic-level differences present a clear opportunity for targeted interventions in primary care, such as standardizing screening workflows across health systems. Provider volume was positively associated with the frequency of visual function tests. However, this should be interpreted with caution due to the limited number of clinics and insufficient variability, which may lead to unstable inferences.

Regarding patient-level factors associated with visual function tests, the reported association between age and visual function tests in this study is consistent with prior literature.^29–31^ Older children (i.e., age 5 or 6 vs 3-years) had higher testability or success with visual function tests.^29–31^ Older children are more likely to receive a visual acuity test because they are usually more cooperative than younger children due to longer attention spans and less anxiety around strangers and unfamiliar devices such as photoscreeners.^29–31^ Also, older children have larger pupils than younger children, making it easier for photoscreeners to capture images.^32^ Regarding red reflex assessment, children aged 3 and 4 were more likely to receive it than those aged 5. Also, female and Multiracial children compared to non-Hispanic white children were more likely to get red reflex assessments. To the best of our knowledge, there are no peer-reviewed studies comparing sex, age, race, or ethnicity, and the receipt of red reflex assessments in pediatric primary care. However, 96% of pediatricians in Ontario, Canada who responded to a survey reported using the red reflex assessment in children under 3 years, but only 29% reported assessing red reflex in children over 3 years.^33^ This may be due to decreased provider concern for retinoblastoma and related pathologies.

Strengths of this study include its novelty in describing variability in vision test rates between clinics within the same health system. Unlike prior studies reporting variability solely between providers, this study demonstrates that variability persists even within the same health system. Additionally, while other studies focused on pediatric primary care clinics and relied on self-reported surveys and interviews, which are subject to bias, this study describes vision screening using objective, real-world data from EHRs thereby enhancing reliability and objectivity.^19,20,23,28^ This is the first study, to the best of our knowledge, to use a retrospective chart review methodology to describe vision screening processes in primary care.

Despite its strengths, this study has limitations. First, this study is limited to three representative pediatric primary care clinics within one university health care system to allow for manual, dual-reviewed data extraction to ensure data accuracy and the capture of unstructured documentation of vision screening in clinical notes that a large-scale, automated EHR pull would miss. Despite the lack of generalizability, study results were similar to studies from other parts of the US, particularly related to age differences in the receipt of vision screening tests.^19,24,28^ Another limitation is the existence of structured EHR macros in the EHR that automatically document an exam, such as red reflex assessments, as completed or not, regardless of whether the exam was actually performed. This may result in an inaccurate enumeration of red reflex assessments. Similarly, the choice of a manual data abstraction strategy increased the risk of manual abstraction inaccuracies. Inaccuracies were mitigated through independent dual review and PI (AA) audit of randomly selected cases. Lastly, data could not be collected on all other potential workflow variables that could be associated with the study outcomes for an deeper assessment of the causes of clinic-level variation.

The high levels of variability in vision tests demonstrated in this study warrant further investigation. Larger multi-clinic studies and qualitative research are needed to identify targetable drivers influencing the delivery of vision-related preventive services. For this to be possible, standardizing screening outcomes in the EHR is essential for surveillance, and the evaluation of interventions. Potential interventions to improve vision screening rates and surveillance include embedding flowsheets in EHRs, and introducing clinical decision support (e.g., templates, reminders, referral orders) supplemented by provider training. Evidence from an Ohio quality improvement collaborative demonstrated that structured efforts can markedly improve screening rates, increasing attempts from 18% to 87%.^24^ Similar tailored interventions could enhance uptake of vision tests and adherence to guidelines.

## Conclusion

About three in five children aged 3 to 5 years received visual function test, while one in five received a red reflex assessment during well-child visits in three pediatric primary care clinics in a university health care system. Clinic and patient factors influenced the likelihood of these assessments. Future research should explore reasons for variability in primary care vision, including qualitative studies with pediatric providers and support staff. Insights from this and subsequent work can guide tailored interventions, such as clinical decision support and provider training, to improve adherence to vision test recommendations and support early identification and intervention for children with vision disorders.

*Declaration of generative AI and AI-assisted technologies: During the preparation of this work the author(s) used the University of Utah’s licensed version of Microsoft Copilot (GPT-4 powered, enterprise-secured, and integrated into Microsoft 365 services) to revise, synthesize, and paraphrase text to enhance the clarity and readability (introduction, and portions of the methods and discussion). After using this tool/service, the author(s) reviewed and edited the content as needed and take(s) full responsibility for the content of the publication*.

## Data Availability

All data produced in the present study are available upon reasonable request to the authors

## Acknowledgements

The authors would like to acknowledge JJ Horns, PhD, for providing statistical consultation for the multivariate analyses presented in this manuscript

## Conflict of Interest

The authors have no conflicts of interest relevant to this article to disclose

## Contributors Statement

Afua Asare conceptualized and designed the study, acquired data, analyzed and interpreted data, and drafted the article. Giovani Robles, Olaoluwa Omotowa, Jessie Montgomery, acquired data, interpreted data and revised it critically for important intellectual content. Bryce Baugh acquired data and revised it critically for important intellectual content. Brian Stagg, JD Smith, Guilherme Del Fiol, Melissa Watt, Eugenie Hartmann, Michelle Hribar, and Carole Stipelman made substantial contributions to conceptualizing and designing the study, interpreting data and revising it critically for important intellectual content. All authors approved the final manuscript as submitted and agree to be accountable for all aspects of the work.

## Funding/Support

This work was supported by the Intermountain Foundation at Primary Children’s Early Career Development Award, the National Institutes of Health Core Grant (EY014800), and an Unrestricted Grant from Research to Prevent Blindness, New York, NY, to the Department of Ophthalmology & Visual Sciences, Spencer Fox Eccles School of Medicine at the University of Utah. The opinions, results, and conclusions reported in this paper are those of the authors and are independent of the funding sources. Funders had no role in the design and conduct of the study.

## Notes

### Competing Interest Statement

The authors have declared no competing interest.

### Author Declarations

IRB of the University of Utah waived ethical approval for this work

