## Supplemental Materials for "Adherence to Red Reflex and Vision Screening Recommendations: A Deep Dive into Primary Care Implementation Gaps"

**SDC, Materials and Methods**

Description of the differences in workflow and populations in the University Health System

Clinic workflow differs between academic and non-academic sites. Academic clinics (e.g., Clinic A and Clinic C) typically schedule 20-minute appointments to accommodate teaching, whereas non-academic clinics (e.g., Clinic B) follow a private practice model with shorter 15-minute visits. Vision screening practices also vary within clinic types. For instance, Clinic C conducts visual acuity testing for children aged 5 years and uses instrument-based screening for younger children, or for children aged 5 and older if cooperation is limited. Clinic A performs only automated instrument-based screening for children aged 3 to 5 years. In contrast, Clinic B relies exclusively on visual acuity testing for children over age 3 and does not use instrument-based tools.

Clinics in the university health care system also differ in the patient populations they serve. Clinic C has the highest racial and ethnic diversity and cares for a larger population of Medicaid and self-pay patients than other clinics in the university health system. In contrast, Clinic A serves the most medically complex patients within the health system. Clinic B serves a more general pediatric population with fewer medically complex cases and less racial and ethnic diversity than Clinic C.

Identifying visual function and red reflex assessments in patient EHRs

The American Academy of Pediatrics recommends annual visual function testing from ages 3 to 5 and red reflex assessments at every well-child visit.^5^ Therefore, vision screening data were collected from EHRs of 3, 4, and 5-year well-child visits. Data abstraction was performed by two independent reviewers per clinic (six total) using a REDCAP survey hosted at the University of Utah, and trained by the lead author (A.A) .^36,37^ Quality checks included a secondary review of half the abstracted data by each of two additional reviewers, resolution of discrepancies, and an audit of a random 10% by the lead author. The lead author also met every two weeks with reviewers to address challenges throughout the data abstraction process. Demographic data (age, sex, race/ethnicity, health insurance, zip code, clinic, and provider counts) were abstracted from the Enterprise Data Warehouse by a data engineer. The entire record for the visit was reviewed, with particular attention to progress notes. The bulk of data on visual function and red reflex assessments was found under objective findings in the chart; specifically, in the ‘*Hearing and Vision Screening*’ Smartform and ‘*Eyes*’ sections, respectively. Also reviewed was ‘*Nursing Notes’*, which often had a summary of the visual function test, and subjective findings under the ‘*Vision/Hearing/Dental*’ section for any patient complaints regarding their vision. When the red reflex was performed, it was recorded as either of the two statements: ‘*Red reflex present bilaterally*’ or the abbreviation ‘*RR+bil*’.

**SDC, Table 1. Vision Screening Tests Discussed in the Study for Children Aged 3 to 5 Years in Pediatric Primary Care**

|  | **Visual Acuity Test ^a^** | **Instrument-based Test^a^** | **Red Reflex (Bruckner) Test ^8^ ^b^** |
| --- | --- | --- | --- |
| **Tool(s)** | HOTV or LEA symbols with lines around each optotype (‘crowding effect’) | Photoscreeners e.g., Plusoptix, Spot Vision Screener, etc. | Direct ophthalmoscope |
| **Purpose** | To measure the clarity or sharpness of vision | To detect eye disorders that may result in poor vision | To identify opacities in the ocular media (optical pathway from the cornea to the retina) and asymmetry in the red reflex in both eyes |
| **Procedure** | Each eye is tested one at a time. The eye not being tested is occluded ( patch or tape). The threshold line or critical line evaluation method is used to determine suboptimal vision. Critical line evaluates children with an age-specific line of chart optotype(s). This method is rapid and ideal for younger children because of its rapid nature.^38^ Visual acuity is the gold standard test once children can identify optotypes (usually age 4 and up, unless there is a developmental delay or disability).^5,38,39^ | Photoscreener is placed in front of the child, who looks at the device briefly while it captures a photo. The photo is then analyzed automatically to detect risk factor conditions for amblyopia. Photoscreening is appropriate for children 18 months old or older. Visual acuity is the gold standard test once children can identify optotypes (usually age 4 and up, unless there is a developmental delay or disability).^5,38,39^ | Examiner holds a direct ophthalmoscope to the child’s eye in a darkened room. Light from the ophthalmoscope is projected onto both eyes simultaneously from about 18 inches away. |
| **Duration** | Critical line: 2-5 minutes, Threshold: 5-10 minutes.^40^ | Under a minute.^41^ | Less than a minute^42^ |
| **Disorders detected** | Refractive error, amblyopia, strabismus, and other vision-threatening conditions. | Refractive error, amblyopia risk factors, strabismus, and other vision-threatening conditions. | Cataracts, glaucoma, retinoblastoma, retinal abnormalities, systemic diseases with ocular manifestations, high refractive error |
| **Normal test** | Child is able to appropriately read the age-specific line of optotype | Magnitude of the error below age-specific thresholds recommended by AAP/AAPOS.^43^ | Red reflex should emanate from both eyes that is symmetrical and with no dark spots |
| **Referrals** | - Child unable to read the age-specific line of optotype. - Child unable to complete the testing (not because they are young, developmentally delayed or have a disability) | - Magnitude of the error above age-specific thresholds recommended by AAP/AAPOS.^43^ - Device is working but unable to complete testing for child. | - Dark spots in the red reflex that remain with blinking, markedly diminished reflex, the presence of a white reflex, or asymmetry of the reflexes (Bruckner reflex). - Regardless of the result of red reflex, children with a positive family history of retinoblastoma, cataracts, glaucoma or retinal abnormalities should also be referred |

^a^ Test of visual function ^b^ Test to determine ocular risks; AAP – American Academy of Pediatrics; AAPOS – American Academy of Pediatric Ophthalmology and Strabismus; Optotype – figures or a selection for distinct letters formatted on chart lines or presented singly on individual charts.^38^

**SDC, Table 2. Provider count across the three pediatric primary care clinics that conducted well-child visits for the year 2022**

| **Personnel n(%)** | **Total (n=63)** | **Well-Child Visit Provider Counts per Clinic** | | | ***P*** |
| --- | --- | --- | --- | --- | --- |
|  |  | **A (n=29)** | **B (n=11)** | **C (n=29)** |  |
| **Attendings n(%)** | 53 | 24 (45.3) | 9 (17.0) | 20 (37.7) | **0.27** |
| **Residents n(%)** | 10 | 5 (0.50) | 0 (0.00) | 5 (50.0) |  |
| **Advanced Practice Clinicians (APC) ^a^** | 2 | 0 (0.0) | 1 (50.0) | 1 (50.0) |  |
| **Allied Health Professionals (AHP) ^b^** | 4 | 0 (0.0) | 1 (25.0) | 3 (75.0) |  |

Chi-square statistic = 7.55, Degrees of freedom = 6, Cramer’s V = 0.234 (small effect between distribution of personnel across the 3 clinics). ^a^ APC includes nurse practitioners and physician assistants; ^b^ AHP includes medical assistants

**SDC, Table 3. Provider count across the three pediatric primary care clinics for the year 2022 (regardless of whether they performed well-child visits or not)**

| **Personnel n(%)** | **Total* (n=133)** | **Provider Counts per Clinic** | | | ***P*** |
| --- | --- | --- | --- | --- | --- |
|  |  | **A (n=65)** | **B (n=54)** | **C (n=37)** |  |
| **Attendings n(%)** | 93 | 46 (49.5) | 44 (47.3) | 25 (26.9) | **<.001** |
| **Residents n(%)** | 25 | 17 (68.0) | 0 (0.00) | 8 (32.0) |  |
| **Advanced Practice Clinicians (APC) ^a^** | 10 | 2 (20.0) | 8 (80.0) | 1 (10.0) |  |
| **Allied Health Professionals (AHP) ^b^** | 5 | 0 (0.00) | 2 (40.00) | 3 (60.00) |  |

Chi-square statistic = 26.21, Degrees of freedom = 6, Cramer’s V = 0.290 (moderate effect between distribution of personnel across the 3 clinics). ^a^ APC includes nurse practitioners and physician assistants; ^b^ AHP includes medical assistants
